# LightOCT: Exploring the depth for Retinal disease detection

**DOI:** 10.1101/2021.11.16.21266390

**Authors:** Amandeep Kaur, Vinayak Singh, Gargi Chakraverty, Dimple Sethi

## Abstract

With the advancement in technology and computation capabilities, identifying retinal damage through state-of-the-art CNNs architectures has led to the speedy and precise diagnosis, thus inhibiting further disease development. In this study, we focus on the classification of retinal damage caused by diabetic macular edema (DME), Choroidal Neovascularization (CNV),DRUSEN, and NORMAL in optical coherence tomography (OCT) images. The emphasis of our experiment is to investigate the component of depth in the neural network architecture. We introduce a shallow convolution neural network - LightOCT, outperforming the other deep model configurations, with the lowest value of LVCEL and highest accuracy (+98% in each class). Next, we experimented to find the best fit optimizer for LightOCT. The results proved that the combination of LightOCT and Adam gave the most optimal results. Finally, we compare our approach with transfer learning models, and LightOCT outperforms the state-of-the-art models in terms of computational cost, least training time and gives comparable results in the criteria of accuracy. We would like to focus our future efforts on improving the accuracy metrics using shallow models, such that the trade-off between training time and accuracy is reduced..

## 1 Introduction

In the domain of healthcare, several imaging methods are deployed to scan the patient’s modalities. In the era of deep learning and computer vision-based methods augmenting the efforts of radiologists, this domain is yet to utilize the full potential of the state-of-the-art proposed techniques in clinical settings. Non-invasive imaging methods like optical coherence tomography (OCT) have shown resolute images of retinal structures. Retinal image study plays a vital role in identifying and further classifying the damage in the retina. OCT images can penetrate through and detect the presence of edema, disruptions in the vasculature of the retina, or the collection of dangerous fluids, which could lead to loss of vision if not treated in the anticipated time. With the advancement in technology and computation powers, identifying retinal damage through state-of-the-art CNNs architectures has led to early and accurate diagnosis, thus preventing further disease development. We focus on the classification of retinal damage caused by detecting diabetic macular edema (DME), Choroidal Neovascularization (CNV),DRUSEN, and NORMAL in optical coherence tomography (OCT) image. We propose LightOCT, a lightweight architecture that can classify the OCT images accurately with their corresponding labels with 98% accuracy in each class. Compared to all other architectures, LightOCT had the least loss, the highest accuracy, and the shortest training time.

The main contributions of this paper are as follows:

- We propose a shallow convolution neural network - LightOCT, for the classification of retinal diseases, with regularization techniques to accelerate convergence and improve accuracy. LightOCT outperformed the other deep model configurations, with the lowest value of Least Validation Cross-Entropy Loss(LVCEL) as 0.2018 and the highest accuracy for each class.
- We experimented to find the best fit optimizer for LightOCT. Adam and RMSProp give comparable results, but the F1 score of the former is better than RMSProp. Thus, the combination of Adam and LightOCT is selected for analysis.
- Next, a comparative analysis of the state-of-the-art transfer learning models and our architecture is conducted. Our model proves to be computationally efficient and requires the least training time.

## 2 Related Work

Lemaître et al. [1] presented a technique for DME detection based on local binary pattern feature extraction from OCT images and a bag of words model, with an 81.2% sensitivity and a 93.7% specificity. The effects of preprocessing, on the other hand, vary depending on the classifier and feature configuration. Gulshan et al. [2] used deep learning to detect diabetic retinopathy and DME, achieving a sensitivity of 90.3% and specificity of 98.1% on the EyePACS-1 dataset and 87.0% sensitivity and 98.5% specificity on the Messidor-2 dataset. However, they have the disadvantage of being computationally expensive.

Alsaih et al. [3]used a support vector machine(SVM) to categorize normal retina and DME with an 87.5% specificity and sensitivity for experimentation on a proprietary dataset. But the limitation is the relatively small dataset used for analysis. Karri et al.[4] proposed a transfer learning based on an Inception network to categorize retinal OCT images into AMD, DME, and Normal with a mean prediction accuracy of 89%, 86%, and 99% respectively. Accuracy should not be recognized as the sole evaluation metric. The paper emphasized only two retinal diseases, but in clinical settings, multi-categorical abnormalities have more commonplace. For many medical imaging tasks, machine learning techniques have given state-of-the-art results. Hussain et al. [5]propose to use a random forest classifier for classifying healthy OCT images from infected ones with a mean accuracy of 96%. However, in the presence of severe pathologies, the model may fail to detect the retinal layers.

Srinivasan et al.[6] used histogram of oriented gradients (HOG) descriptors and SVM classifiers for detecting AMD, DME, and normal retina, attaining accuracies nearly 100%, 100%, and 88.87% respectively. One major drawback of such approaches is their reliance on features/markings defined by ophthalmologists, which may lead to delayed and inefficient generalization, further questioning the feasibility of this approach on larger datasets. Ten et al.[7] using a ten-fold cross-validation strategy and deep convolution neural network(CNN), detected AMD with a mean accuracy of 95.45%, the sensitivity of 96.43%, and specificity of 93.75%. Lu et al. [8] proposed to detect and differentiates multi-categorical abnormalities from OCT images automatically; in their work, the image dataset is randomly divided into train, validation, and test at the image level, which may lead to a bias, as multiple images from the same eye could be present in different partitions. Though their mean accuracy is 95.9%, with 97.3% specificity and 94.0% sensitivity, the model’s performance is partial. Li et al.[9] use the VGG-16 network to obtain the classification results on AMD and DME, with 98.6% accuracy, a 97.8% sensitivity, a 99.4% specificity, but since the validation set of 1000 images was used as the testing set, the results of the model may be biased. Kermany et al. [10] used the Inception V3 network to report an accuracy of 96.6%, with a sensitivity of 97.8%, and specificity of 97.4%, by performing occlusion, they tried to detect the areas contributing richly to the neural net’s final prediction. But the model suffers a vanishing gradient problem, which could affect the results. Fauw et al. [11] developed an ensemble of segmentation(U-net) and classification networks(DenseNet) to detect OCT scans. The accuracy of testing dataset 1 was 94.5%, and on the testing dataset 2 was 96.6%. The major drawback was the time complexity. Table 1 summarizes the existing methods.

**Table 1.** Literature review: Details and results of the existing methods for retinal disease detection using OCT

| Ref | Method | Results | Limitation |
| --- | --- | --- | --- |
| [1] | BoW and local binary pattern feature | Sensitivity of 81.2%, specificity of 93.7% for DME detection | Effects of pre-processing differ depending on the classifier and feature configuration |
| [2] | Deep learning | Sensitivity of 90.3% and specificity 98.1% on the EyePACS-1 dataset and 87.0% sensitivity and 98.5% specificity on the Messidor-2 dataset | Computationally expensive |
| [3] | Support Vector Machine(SVM) | Sensitivity and specificity of 87.5% to categorize normal retina and DME on a proprietary dataset | Small dataset used for analysis |
| [4] | Transfer learning based on an Inception network | AMD, DME, and Normal with a mean prediction accuracy of 89%, 86%, and 99% respectively | Paper emphasized only two retinal diseases |
| [5] | Random forest classifier | Mean accuracy 96% | In severe pathologies, detection of the retinal layers may fail |
| [6] | Histogram of oriented gradients (HOG) descriptors and SVM classifiers | Attaining accuracies for Detecting of AMD, DME, and normal retina as 100%, 100%, and 88.87% respectively | Delayed and inefficient generalization |
| [7] | Deep Convolution Neural Network(CNN) | Detected AMD with mean accuracy of 95.45%, sensitivity of 96.43%, and specificity of 93.75% | Small dataset used for analysis |
| [8] | Convolution Neural Network(CNN) - ResNet 101 | Mean accuracy is 95.9%, with 97.3% specificity and 94.0% sensitivity | Performance of the model is partial as tested on validation test set |
| [9] | Convolution Neural Network(CNN) - VGG 16 | Classification results on AMD and DME, with 98.6% accuracy, a 97.8% sensitivity, a 99.4% specificity | Performance of the model is partial as tested on validation test set |
| [10] | Inception V3 network, performed occlusion | Accuracy of 96.6%, with a sensitivity of 97.8%, and a specificity of 97.4% | Model suffers a vanishing gradient problem |
| [11] | Ensemble of U-net and DenseNet | Accuracies on the testing dataset 1 (Device Type 1) was 94.5% and on the testing dataset 2 (Device Type 2) was 96.6% | Time complexity |

## 3 Methodology

### 3.1 Convolution Neural Networks (CNNs)

Convolutional Neural Networks (CNNs) have proven to be the best-performing visual data architectures, particularly in the field of computer vision. CNNs have demonstrated exceptional performance in image classification, segmentation, detection, and related tasks, according to Cirega et al.[12]. This technology is being used outside of academia, with large corporations (Microsoft, Meta, Google, and ATT) having research groups actively investigating the benefits of CNNs. CNN’s most notable feature is its ability to use both spatial and temporal correlation data. CNN learns through the back-propagation algorithm during the training phase, with the architecture containing alternate layers of convolution and pooling, followed by the final layers, which are fully connected, as introduced by LeCun et al. [13].

The following subsections detail the internal structures of various layers in the CNN.

#### 3.1.1 Convolutional Layer

The convolutional layer is made up of a set of kernels with each neuron acting as a kernel.[14]. These kernels further divide the image into small patches called receptive fields. Finally, features are extracted from these fields. The kernel convolves through the image with weights by multiplying its elements with the corresponding parts of the receptive field, Bouvrie et al.[15] as described in equation 1 as,

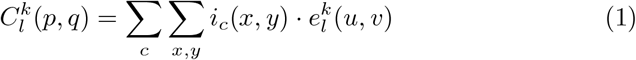

where *i*_*c*_(*x, y*) is an element of the input image tensor *I*_*C*_, which is element wise multiplied by 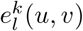 index of the *k* th convolutional kernel *k*_*l*_ of the *l* th layer. In contrast, the output feature-map of the *k* th convolutional operation can be written as 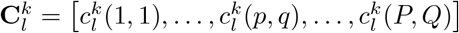

#### 3.1.2 Pooling layer

The pooling method facilitates the extraction of a set of features that are insensitive to translational shifts and small distortions, Huang et al. [16]

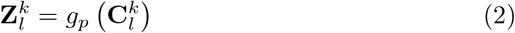

Equation 2 shows the pooling operation in which 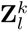 represents the pooled featuremap of *l* th layer for *k* th input feature-map 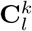, whereas *g*_*p*_(.) defines the type of pooling operation.

#### 3.1.3 Activation function

The activation function serves as a decision function and aids in the learning of complex patterns.

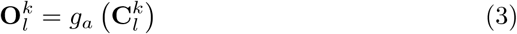

In the preceding equation 3, 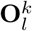 is an output of a convolution, which is assigned to activation function *g*_*a*_(.) that adds non-linearity and returns a transformed output 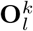 for *l* th layer.

#### 3.1.4 Batch normalisation

Batch normalisation is used to solve problems caused by internal covariance shifts in feature-maps.

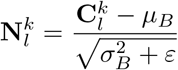

Where 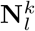 represents normalized feature-map, 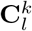 is the input feature-map, *µ*_*B*_ and 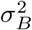 depict mean and variance of a feature-map for a mini batch respectively. In order to avoid division by zero, *ε* is added for numerical stability. According to Szegedy et al. [17], batch normalisation unifies the distribution of feature-map values by setting the distribution to zero mean and unit variance.

### 3.2 Optimizer

Optimizers are responsible for adjusting the characteristics of a neural network, such as hyperparameters like learning rate and weight, in order to reduce loss. We experimented to identify the best fit optimizer for the proposed design, which turned out to be LightOCT. For our trials, we used a variety of optimizers, as indicated below. in Fig 2.

**Fig. 1.**
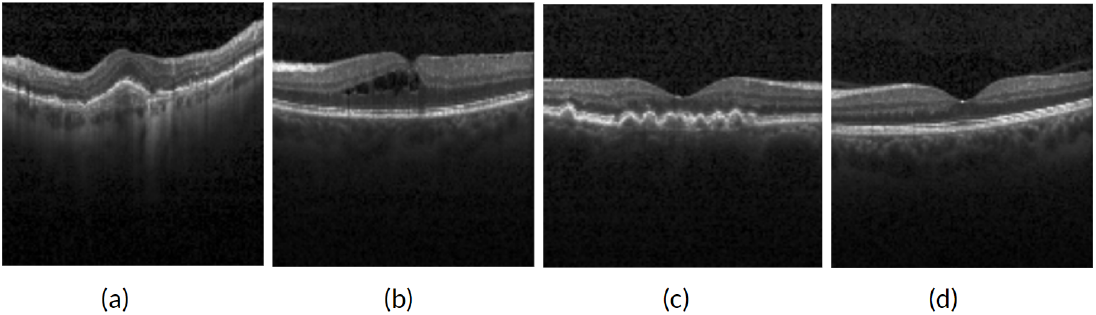
Representative optical coherence tomography (OCT) images. (a) CNV - Choroidal neovascularization, (b) DME - Diabetic Macular Edema, (c) DRUSEN, (d) Normal Retina

**Fig. 2.**
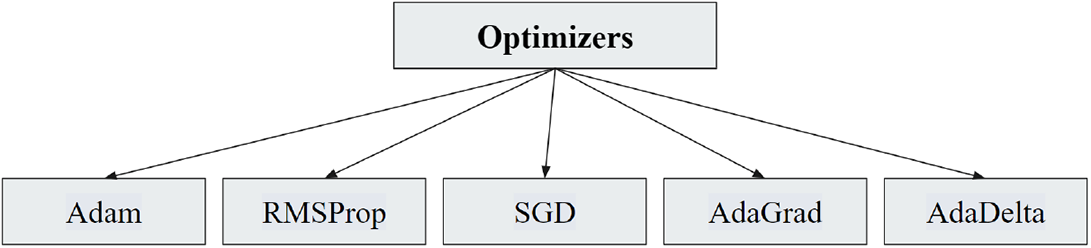
Various optimizers used

#### Adam

Kingma et al.[18] proposed Adam stands for adaptive moment estimation. Instead of the classical gradient descent procedure, it is based on the adaptive analysis of first and second-order moments. It combines the benefits of two other stochastic gradient descent extensions: SGD with momentum and

RMSProp, resulting in an optimization algorithm that can handle sparse gradients on noisy problems.

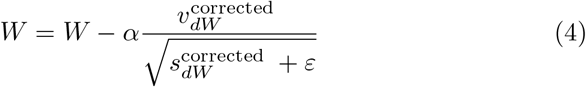

Here W is the weight of matrix, *α* is the learning rate, *v*_*dW*_ is the exponentially weighted average of past gradients and *s*_*dW*_ is the exponentially weighted average of past squares of gradients with *ε* as a small value to avoid dividing by zero.

In our experiment, Adam gives the least LVCEL value with maximum MVA with LightOCT, as seen in Table 4.

#### RMSProp

RMSprop, which stands for Root Mean Square Propagation, was proposed by Geoff Hinton. It is a mini-batch learning optimization technique based on gradients. It solves the problem of vanishing gradients in very complex neural network functions by normalising the gradient with a moving average of squared gradients. RMS-Prop is a combination of momentum and AdaGrad.

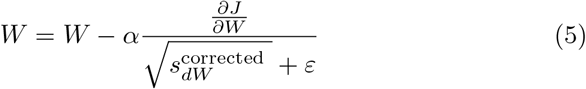

Here, W is the weight tensor,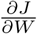 refers to the cost gradient wrt to current W, and *s*_*dW*_ is the exponentially weighted average of past squares of gradients with *ε* as a small value to avoid dividing by zero.

With the proposed model, RMSProp gave the second highest MVA and second least LVCEL value. The results are comparable with Adam, but the F1 score for Adam is better than RMSProp. Refer Table 4.

#### SGD

Iterative optimization technique stochastic gradient descent (SGD) provides a stochastic approximation. In high-dimensional optimization problems, it is used to reduce computational burden and achieve faster iterations in exchange for a lower convergence rate.

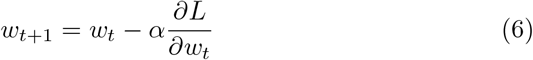

Here, 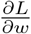 refers the gradient of *L*, the loss function to be minimised, w.r.t. to *w, α* as the learning rate, *t*, the time step and *w* is the weight/parameter which needs to be updated where the subscript *t* indexes the weight at time step *t*.

SGD result on MVA is close to 91% but it’s LVCEL value is higher than the former two optimizers.

#### AdaGrad

Adagrad (adaptive gradient) is an optimizer with parameter-specific learning rates. Learning rates are proportional to how frequently a parameter is updated during training and are inversely proportional to the sum of the squares of all the parameter’s previous gradients.

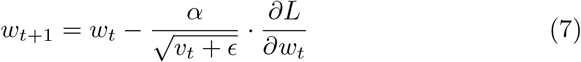

where

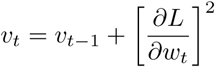

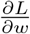 is the gradient of *L*, the loss function to minimise, w.r.t. to *w, α* is the learning rate, *t* implies the time step, *w* is the weight/parameter that we want to update, where the subscript *t* indexes the weight at time step *t, v* is the cumulative sum of current and past squared gradients (up to time *t*), initialised to *o*, and *ε* is a small floating point value that ensures we never have to divide by zero. AdaGrad achieves an MVA of only 83 percent with a twofold increase in LVCEL value over RMSProp.

#### AdaDelta

Matthew D. Zeiler et al. [19] proposed Adadelta, an optimization algorithm designed to address two shortcomings of the Adagrad method: the decaying learning rate problem during training and the need to manually select a global learning rate. Instead of accumulating all previous squared gradients, Adadelta adjusts the learning rate by limiting the accumulated previous gradients with a fixed-size moving window. Adadelta’s equation is written as

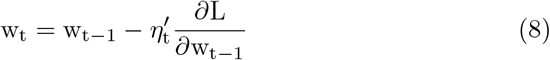

where w is the weight tensor,

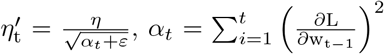 summation of gradient square and *ε* is a small + ve number to avoid divisibilty by 0.

AdaDelta managed to reach an MVA of 71% with a 4 times increase in value of LVCEL when compared to Adam.

#### 3.2.1 Loss Function

Loss functions can be defined as a prediction error of neural networks. Gradients are used to update the weights of a neural net, and the Loss is used to calculate them. For our experiment, we tried two loss functions - sparse categorical cross-entropy and Square-hinge loss.

##### Sparse categorical cross-entropy

When adjusting model weights during training, cross-entropy loss is used. It is mathematically defined as:

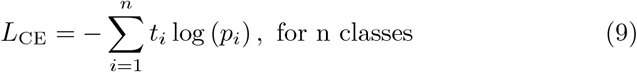

where *t*_*i*_ is the truth label and *p*_*i*_ is the Softmax probability for the *i*^th^ class.

Sparse categorical cross-entropy has the same loss function as defined in the preceding equation and is used when the targets are integers rather than categorical vectors.

##### Square-hinge

The hinge loss function, which was developed primarily with Support Vector Machine (SVM) models, encourages examples to have the correct sign and assigns more error when the predicted and actual class values differ in sign. It smoothes the surface of the error function, making it easier to work with numerically. It has several extensions, including Squared hinge loss, which computes the square of the score hinge loss. The target variable, like the hinge loss function, must be modified to have values in the range [-1, 1]. Hinge loss of the prediction *y* is defined as:

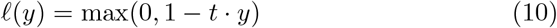

Here *y* should be the ‘raw’ output of the classifier’s decision function,not the predicted class label. For squared Hinge loss we calculate the square of the above score hinge loss.

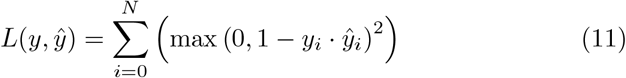

where is the ŷ the predicted value and y is either 1 or -1

### 3.3 Proposed Approach

We implemented various configured CNN architectures in which base models were classified into three categories - LightOCT, MediumOCT, and HeavyOCT, the configurations of those are listed in Table 2. Compared to the other two architectures, LightOCT had the fewest parameters, resulting in less training time. As a result of the relationship, we can conclude that parameters are directly proportional to training time. Because LightOCT had the fewest parameters, the proposed model also reduced training time. For training and selecting the best-configured architecture, the Adam optimizer was used as a base optimizer. HeavyOCT had 2.4 million parameters, which required more training time. We also tried regularization techniques - batch normalization and dropouts for the models. Adam optimizer was used for the initial implementation, and we found the optimal learning rate as 1.00*e* − 05.

**Table 2.** Configuration of CNN architectures

| Model Name | CL<sup>1</sup> | AL<sup>2</sup> | Parameters | Optimizer Used | Learning Rate of Optimizer | BN<sup>3</sup> | DO<sup>4</sup> | FD<sup>5</sup> | DL<sup>6</sup> | KS<sup>7</sup> | PS<sup>8</sup> |
| --- | --- | --- | --- | --- | --- | --- | --- | --- | --- | --- | --- |
| LightOCT | 3 | 2 | 240,820 | Adam | 1.00E-05 | ✓ | × | (32,64,128) | {16,32} | (3,3,3) | (2,2,2) |
| MediumOCT | 4 | 3 | 426,388 | Adam | 1.00E-05 | × | ✓ | (32,64,64,128) | {16,64,128} | (3,3,3,6) | (2,2,2,4) |
| HeavyOCT | 6 | 6 | 2,406,788 | Adam | 1.00E-05 | ✓ | ✓ | (32,32,64,64,128,128) | {16,16,64,128,256,256} | (3,3,3,6,9,9) | (2,2,2,2,2,2) |

<sup>1</sup>Convolution Layer, <sup>2</sup>Artificial Layer, <sup>3</sup>Batch Normalisation, <sup>4</sup>Dropout, <sup>5</sup>Feature Detected, <sup>6</sup>Dense Layer, <sup>7</sup>Kernel Size, <sup>8</sup>Pooling Size

We selected LightOCT as our final model for further analysis. In LightOCT, we used batch normalization to accelerate convergence and improve accuracy.

As seen in Fig. 3, our model consists of 3 convolution layers with varying kernel sizes, followed by a 2 ⨯ 2 max-pooling layer and 2 fully connected dense layers, predicting the output for the 4 classes, with Adam as the optimizer.

**Fig. 3.**
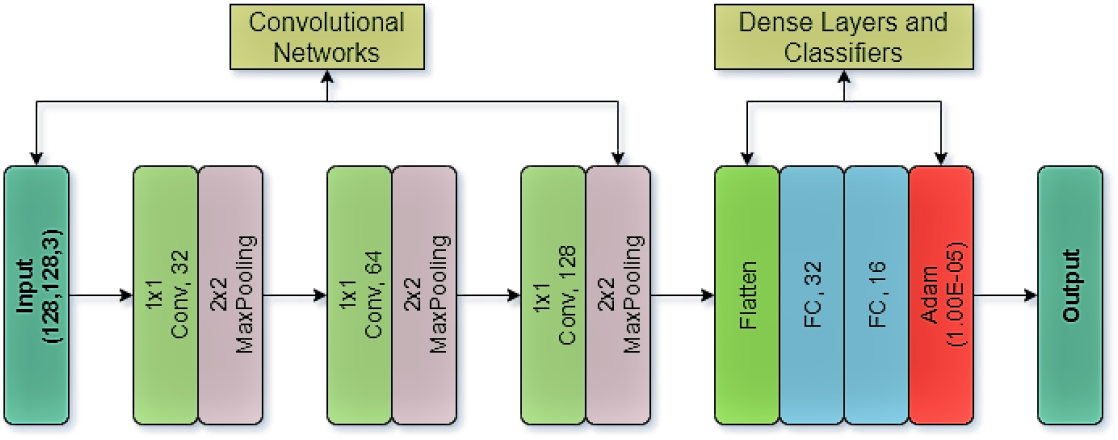
Architecture of proposed model - LightOCT

### 3.4 Comparative analysis with Transfer learning

Transferring learning is an essential technique in deep learning because it produces accurate models in a short period. Rather than starting from scratch, the model in this technique learns from patterns previously used to solve a different problem. We employ pre-trained models that have been trained on benchmark data sets such as ImageNet [20], CIFAR-10 Dataset [21], PASCAL VOC Dataset [22], and others. This section provides an overview of the pre-trained models that were used in our analysis.

#### VGG

Simonyan and Zisserman [23] introduced the revolutionary VGG Network, which is a Spatial Exploitation based CNN. The input to the VGG-based convNet is a 224 ⨯ 224 RGB image. The preprocessing layer subtracts the mean image values (calculated across the entire ImageNet training set) from the RGB image (with pixel values ranging from 0 to 255). These weight layers are applied to the input images after they have been preprocessed. The training images are processed by a convolution layer stack. There are 13 convolutional layers and three fully connected layers in the VGG16 architecture. Instead of large filters, VGG has smaller filters (3 ⨯ 3) with greater depth. It has the same effective receptive field as if only one convolutional layer of 7 ⨯ 7 was used. Another VGGNet variant includes 19 weight layers (16 convolutional layers and three fully connected layers) as well as five pooling layers. Both variations of VGGNet have two fully connected layers with 4096 channels each, followed by another fully connected layer with 1000 channels to predict 1000 labels (VGG19 and VGG16). In the final fully connected layer, the softmax layer is used for classification. Both image classification and localization problems were handled well by VGG. VGG came in second place in the 2014-ILSVRC competition, but it quickly rose to prominence due to its simplicity, homogeneous topology, and increased depth. VGG’s main limitation was the use of 138 million parameters, which made it computationally expensive and difficult to deploy on low-resource systems.

#### ResNet

He et al. [24] introduced ResNet50, which features a four-stage architecture. ResNet is depth-based and multi-path-based CNN. This architecture is widely used; even with many layers, training can be done easily without an increase in error percentage. Another advantage is that by using identity mapping, ResNets aid in addressing the vanishing gradient problem. The input image supplied to the network has height, width as multiples of 32, and a channel width of 3. The input size is typically 224 ⨯ 224 ⨯x 3. To perform initial convolution and max-pooling, this design employs 7 ⨯ 7 and 3 × 3 kernel sizes. The first stage of the network is comprised of three Residual blocks, as illustrated in Fig 4, each with three layers. The kernel sizes for the convolution process in stage 1’s three layers are 64, 64, and 128. The Residual Block’s convolution operation uses a stride of 2, which means that the input size is reduced to half in terms of height and width, but the channel width is doubled. Each stage doubles the width of the channel while the input size gets reduced to half. Deeper networks, such as ResNet 50, ResNet101, and ResNet152, incorporate bottleneck design. A Bottleneck Residual Block is a form of a residual block that creates a bottleneck using 1×1 convolutions. To increase the depth and decrease parameters, we make residual blocks as thin as feasible. For each residual function F, the three layers with 1 ⨯ 1, 3 ⨯3, and 1 ⨯ 1 convolutions are stacked one on top of the other. An Average Pooling layer, followed by a fully connected layer, completes the network. ResNet won the 2015-ILSVRC competition with its 152-layer deep CNN proposal.

**Fig. 4.**
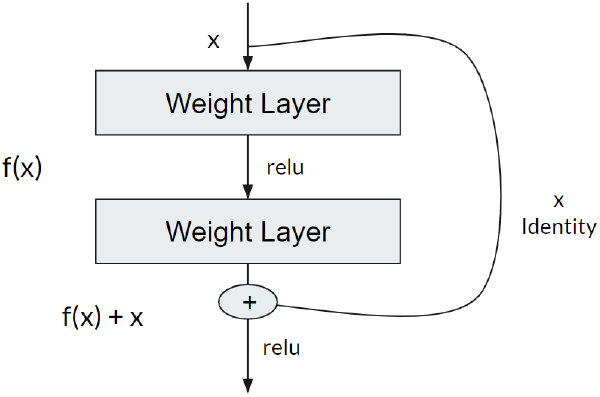
Residual Learning: A building block

#### Inception

The computational efficiency of training more extensive networks was of special interest to the authors of Inception [25]. Inception has width-based multi-connection CNN architecture. Inception modules perform many different transformations over the same input map, concatenating the results into a single output. In other words, Inception does a 5 ⨯ 5 convolutional transformation, a 3 ⨯ 3, and a max-pool for each layer. And the next layer of the model gets to decide if (and how) to use each piece of information. For dimensionality reduction, 1 ⨯ 1 convolutions are used. 1 ⨯ 1 convolutions are used to “ filter” the outputs’ depth to solve the computational bottleneck. When used across multiple channels, a 1 × 1 convolution only looks at one value at a time, but it can extract spatial information and compress it down to a lower dimension. By reducing the number of input maps, the Inception authors were able to stack different layer transformations in parallel, resulting in nets that were both “ deep” (many layers) and “ wide” (many parallel operations).

#### Xception

Chollet introduced Xception, which stands for “ Extreme Inception,” [26]. And, as the title suggests, it takes the principles of Inception to their logical conclusion. Rather than dividing input data into many compressed chunks, Xception maps the spatial correlations for each output channel separately before performing a 1 ⨯ 1 depthwise convolution to capture cross-channel correlation. On the ImageNet dataset, Xception slightly outperforms Inception V3, but vastly outperforms it on a larger image classification dataset with 17,000 classes. It also has the same number of model parameters as Inception, implying that it is more computationally efficient. According to [27], Xception simplifies computation by convolving each feature-map separately across spatial axes, followed by pointwise convolution (1 ⨯ 1 convolutions) to perform cross-channel correlation.

## 4 Experimental Setup

### 4.1 Dataset

The current study made use of 84,495 optical coherence tomography (OCT) images collected by Kermany et al. in [28]labeled from various tertiary hospitals [Beijing Tongren Eye Center, California Retinal Research Foundation, Shiley Eye Institute of the University of California San Diego, Shanghai First People’s Hospital, Medical Center Ophthalmology Associates] between July 1, 2013, and March 1, 2017. Clinical approval has been granted for the data set. Each OCT image was graded using a multi-tiered system. Undergraduate and medical students who had completed an OCT review course completed the initial grading round. Any OCT images with artefacts or poor image resolution were discarded by this group. The first group’s images were approved or rejected by the next tier of four ophthalmologists.

Based on their level of expertise, they detected the presence or absence of various pathologies visible on the OCT. The final tier, which included two retinal specialists with over 20 years of clinical experience, validated the truth labels for each image. A validation subset of 993 OCT images was also verified by two additional ophthalmologist graders to account for human error in grading. The final dataset contains 37,205 class CNV images, 11,348 DME images, 8616 drusen images, and 26,315 standard images. Each class has 242 images in the test data.

## 5 Results and Evaluation

According to Table 3, LightOCT performed exceptionally well, achieving the highest accuracy for each class on test data. LightOCT had a minimum LVCEL of 0.2018 as well. Compared to other architectures, MediumOCT had the lowest validation accuracy while the recall value gained by each class of LightOCT was extremely high. A comparitive analysis of performance between the different configurations tried is shown in the Fig. 5. As a result, we chose LightOCT as the most suitable architecture for further consideration.

**Table 3.** Metric Evaluation of CNN models

| Model Name | MVA | LVCEL | Class | Testing Accuracy | Testing Precision | Testing Recall | Testing F1 Score | Confusion Matrix |  |  |  |
| --- | --- | --- | --- | --- | --- | --- | --- | --- | --- | --- | --- |
| LightOCT | 0.9357 | 0.2018 | CNV | 0.9876 | 0.99 | 0.96 | 0.98 | 240 | 2 | 0 | 0 |
|  |  |  | DME | 0.9907 | 0.97 | 0.99 | 0.98 | 6 | 235 | 1 | 0 |
|  |  |  | DRUSEN | 0.9845 | 0.98 | 0.96 | 0.97 | 4 | 0 | 238 | 0 |
|  |  |  | NORMAL | 0.9897 | 0.96 | 1.0 | 0.98 | 0 | 0 | 10 | 232 |
| MediumOCT | 0.7986 | 0.5583 | CNV | 0.8791 | 0.98 | 0.68 | 0.80 | 236 | 6 | 0 | 0 |
|  |  |  | DME | 0.9008 | 0.72 | 0.86 | 0.78 | 24 | 175 | 3 | 40 |
|  |  |  | DRUSEN | 0.7965 | 0.20 | 0.94 | 0.33 | 87 | 22 | 48 | 85 |
|  |  |  | NORMAL | 0.9897 | 0.96 | 1.0 | 0.98 | 0 | 0 | 10 | 232 |
| HeavyOCT | 0.8421 | 0.4405 | CNV | 0.9566 | 0.98 | 0.87 | 0.92 | 236 | 5 | 1 | 0 |
|  |  |  | DME | 0.9442 | 0.91 | 0.87 | 0.89 | 3 | 221 | 1 | 17 |
|  |  |  | DRUSEN | 0.8874 | 0.56 | 0.99 | 0.71 | 33 | 27 | 135 | 47 |
|  |  |  | NORMAL | 0.9329 | 1.0 | 0.79 | 0.88 | 0 | 1 | 0 | 241 |

**Table 4.** Metric evaluation of LightOCT with different optimizers

| Optimizer | MVA | LVCEL | Class | Testing Accuracy | Testing Precision | Testing Recall | Testing F1 Score | Confusion Matrix |  |  |  |
| --- | --- | --- | --- | --- | --- | --- | --- | --- | --- | --- | --- |
| Adam | 0.9357 | 0.2018 | CNV | 0.9876 | 0.99 | 0.96 | 0.98 | 240 | 2 | 0 | 0 |
|  |  |  | DME | 0.9907 | 0.97 | 0.99 | 0.98 | 6 | 235 | 1 | 0 |
|  |  |  | DRUSEN | 0.9845 | 0.98 | 0.96 | 0.97 | 4 | 0 | 238 | 0 |
|  |  |  | NORMAL | 0.9897 | 0.96 | 1.0 | 0.98 | 0 | 0 | 10 | 232 |
| RMSProp | 0.9248 | 0.2248 | CNV | 0.9421 | 0.98 | 0.82 | 0.89 | 238 | 0 | 0 | 4 |
|  |  |  | DME | 0.8337 | 0.33 | 1.0 | 0.50 | 29 | 81 | 1 | 131 |
|  |  |  | DRUSEN | 0.9587 | 0.84 | 1.0 | 0.91 | 23 | 0 | 203 | 16 |
|  |  |  | NORMAL | 0.8440 | 1.0 | 0.62 | 0.76 | 0 | 0 | 0 | 242 |
| SGD | 0.9079 | 0.2694 | CNV | 0.8122 | 0.69 | 0.61 | 0.65 | 167 | 75 | 0 | 0 |
|  |  |  | DME | 0.5810 | 1.0 | 0.37 | 0.54 | 0 | 242 | 0 | 0 |
|  |  |  | DRUSEN | 0.7750 | 0.10 | 0.96 | 0.19 | 107 | 110 | 25 | 0 |
|  |  |  | NORMAL | 0.7730 | 0.091 | 1.0 | 0.17 | 0 | 220 | 0 | 22 |
| AdaGrad | 0.8384 | 0.4535 | CNV | 0.9132 | 0.98 | 0.75 | 0.85 | 236 | 5 | 1 | 0 |
|  |  |  | DME | 0.9494 | 0.88 | 0.92 | 0.90 | 10 | 212 | 0 | 20 |
|  |  |  | DRUSEN | 0.8688 | 0.48 | 0.98 | 0.65 | 68 | 13 | 117 | 44 |
|  |  |  | NORMAL | 0.9318 | 0.99 | 0.79 | 0.88 | 0 | 1 | 1 | 240 |
| AdaDelta | 0.7112 | 0.8020 | CNV | 0.8347 | 0.96 | 0.61 | 0.74 | 233 | 4 | 0 | 5 |
|  |  |  | DME | 0.8068 | 0.31 | 0.78 | 0.45 | 62 | 76 | 4 | 100 |
|  |  |  | DRUSEN | 0.7655 | 0.079 | 0.83 | 0.14 | 88 | 17 | 19 | 118 |
|  |  |  | NORMAL | 0.7686 | 1.0 | 0.52 | 0.68 | 1 | 0 | 0 | 241 |

**Fig. 5.**
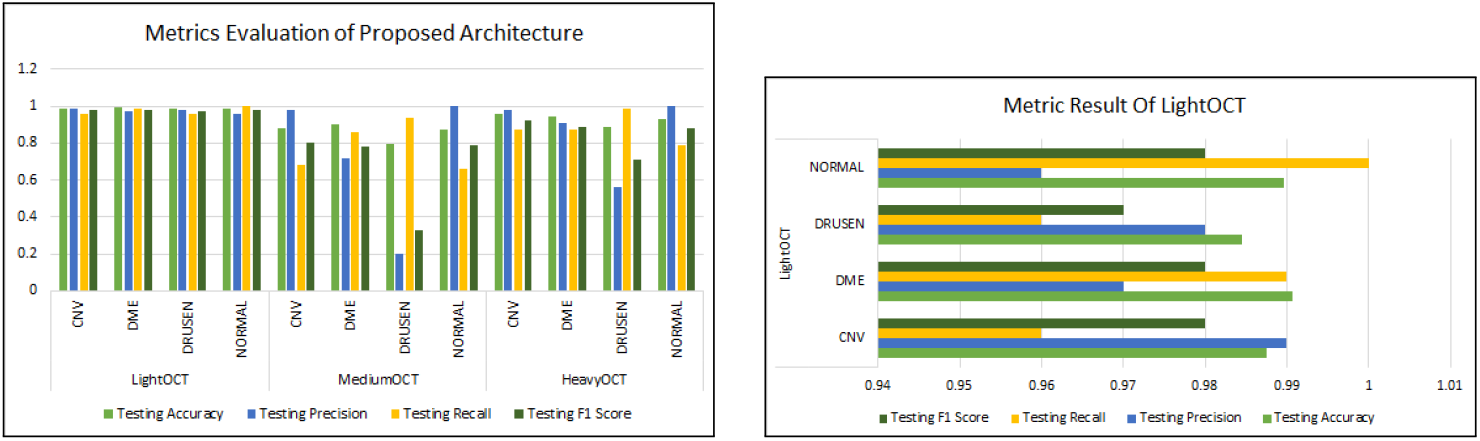
(a) Represents metric evaluation of the proposed architectures, LightOCT, MediumOCT and HeavyOCT, (b) Metric result with the final proposed architecture LightOCT

Next, we tested various optimizers to find the best fit for our LightOCT architecture. Table 4 shows that the results obtained by default Adam were very high when compared to other optimizers. AdaDelta produced inferior results, and LVCEL was more significant with the least value of MVA. As a result, we withdraw from using Adadelta for our LightOCT. RMSProp was a close competitor for Adam, with RMSprop gaining 0.9248 accuracies and Adam trailing by 0.9357. When Adam was used in conjunction with LightOCT, F1 scores in each class were extremely high. We finally chose the combination of Adam and LightOCT to be the best optimal condition for detecting Retinal disease using OCT images. A comparitive analysis of performance between the different optimizers tried with the architecture LightOCT is shown in the Fig. 6.

**Fig. 6.**
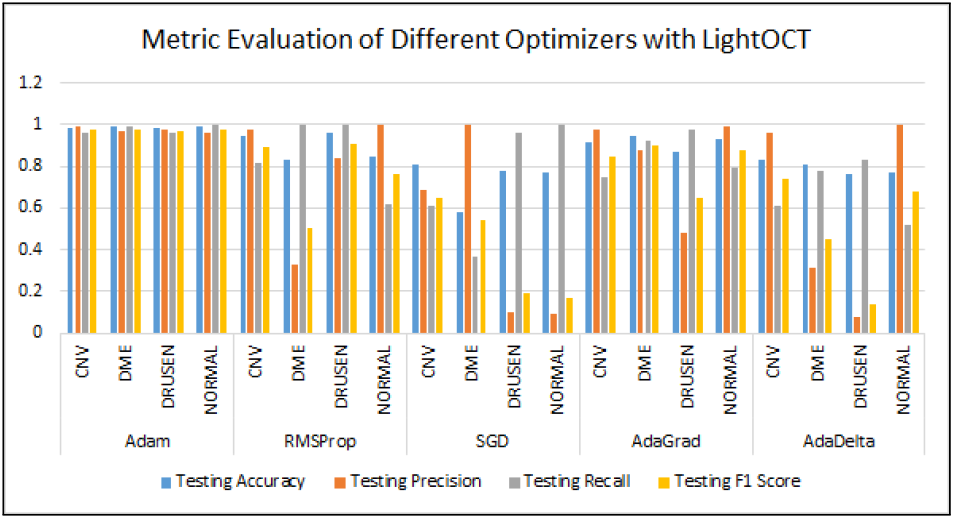
Metric evaluation of Optimizers - Adam, RMSProp, SGD, AdaGrad, AdaDelta with proposed architecture LightOCT

The comparison of our architecture with transfer learning models is shown in Table 5. ResNet50 and ResNet152 outperformed LightOCT in terms of accuracy, but our architecture beats all other transfer learning models when it comes to low computational cost. When compared to other transfer learning models, LightOCT required the least amount of training time. The time required for training by VGG19 and VGG16 was huge, as was the computational cost. The results obtained were less-comparable. Xception had a considerable loss of 2.1132 and a short accuracy gain of 0.6477. The metric evaluation of all transfer learning models is shown in Table 5. Compared to all other architectures as seen in Fig. 7, LightOCT had the least loss, the highest accuracy gained, and the shortest training time. As a result, LightOCT proves to be the most influential architecture for detecting retinal disease using Optical Coherence Tomography images, which is computationally low in cost. Since our model performs efficiently in both time-complexity as well as computational costs, we also provide the activation of the neural network on the four classes, as shown in Fig 8 and Fig 9 to analyze more profoundly into the explainability part of the predictions.

**Table 5.** Comparative analysis of Light with transfer learning models

| Model | MVA | LVCEL | Class | Testing Accuracy | Testing Precision | Testing Recall | Testing F1 Score | Confusion Matrix |  |  |  |
| --- | --- | --- | --- | --- | --- | --- | --- | --- | --- | --- | --- |
| VGG16 | 0.8595 | 0.3665 | CNV | 0.9220 | 0.99 | 0.77 | 0.86 | 242 | 1 | 0 | 1 |
|  |  |  | DME | 0.9435 | 0.79 | 0.98 | 0.87 | 12 | 192 | 1 | 39 |
|  |  |  | DRUSEN | 0.9148 | 0.66 | 0.99 | 0.80 | 62 | 2 | 162 | 18 |
|  |  |  | NORMAL | 0.9405 | 1.0 | 0.81 | 0.89 | 0 | 0 | 0 | 242 |
| VGG19 | 0.7117 | 0.5798 | CNV | 0.8088 | 1.0 | 0.57 | 0.72 | 242 | 1 | 0 | 0 |
|  |  |  | DME | 0.8906 | 0.57 | 0.99 | 0.73 | 24 | 142 | 6 | 76 |
|  |  |  | DRUSEN | 0.8149 | 0.29 | 0.92 | 0.44 | 162 | 0 | 70 | 13 |
|  |  |  | NORMAL | 0.9090 | 1.0 | 0.73 | 0.84 | 0 | 0 | 0 | 242 |
| ResNet50 | 0.9524 | 0.1326 | CNV | 0.9888 | 1.0 | 0.96 | 0.98 | 242 | 0 | 0 | 0 |
|  |  |  | DME | 0.9959 | 0.98 | 1.0 | 0.99 | 4 | 242 | 0 | 0 |
|  |  |  | DRUSEN | 0.9928 | 0.97 | 1.0 | 0.99 | 7 | 0 | 242 | 0 |
|  |  |  | NORMAL | 1.0000 | 1.0 | 1.0 | 1.0 | 0 | 0 | 0 | 242 |
| ResNet101 | 0.9225 | 0.1791 | CNV | 0.9291 | 1.0 | 0.78 | 0.88 | 242 | 0 | 0 | 0 |
|  |  |  | DME | 0.9969 | 0.99 | 1.0 | 0.99 | 2 | 242 | 0 | 1 |
|  |  |  | DRUSEN | 0.9260 | 0.70 | 1.0 | 0.83 | 67 | 0 | 172 | 5 |
|  |  |  | NORMAL | 0.9938 | 1.0 | 0.98 | 0.99 | 0 | 0 | 0 | 242 |
| ResNet152 | 0.9576 | 0.1166 | CNV | 0.9598 | 1.0 | 0.86 | 0.93 | 242 | 1 | 0 | 0 |
|  |  |  | DME | 0.9897 | 0.97 | 0.99 | 0.98 | 7 | 232 | 0 | 1 |
|  |  |  | DRUSEN | 0.9680 | 0.87 | 1.0 | 0.93 | 31 | 0 | 212 | 0 |
|  |  |  | NORMAL | 0.9979 | 1.0 | 1.0 | 1.0 | 0 | 1 | 0 | 242 |
| InceptionV3 | 0.5898 | 2.3727 | CNV | 0.7708 | 0.094 | 0.96 | 0.17 | 23 | 222 | 0 | 0 |
|  |  |  | DME | 0.6012 | 1.0 | 0.38 | 0.56 | 0 | 242 | 0 | 0 |
|  |  |  | DRUSEN | 0.8222 | 0.28 | 1.0 | 0.44 | 1 | 162 | 67 | 10 |
|  |  |  | NORMAL | 0.9856 | 0.99 | 0.96 | 0.97 | 0 | 3 | 0 | 242 |
| Xception | 0.6477 | 2.1132 | CNV | 0.7933 | 0.18 | 0.98 | 0.31 | 45 | 202 | 0 | 0 |
|  |  |  | DME | 0.6527 | 1.0 | 0.42 | 0.59 | 0 | 242 | 0 | 0 |
|  |  |  | DRUSEN | 0.8554 | 0.42 | 1.0 | 0.59 | 0 | 132 | 102 | 9 |
|  |  |  | NORMAL | 0.9837 | 0.97 | 0.96 | 0.97 | 0 | 7 | 0 | 242 |
| LightOCT | 0.9357 | 0.2018 | CNV | 0.9876 | 0.99 | 0.96 | 0.98 | 240 | 2 | 0 | 0 |
|  |  |  | DME | 0.9907 | 0.97 | 0.99 | 0.98 | 6 | 235 | 1 | 0 |
|  |  |  | DRUSEN | 0.9845 | 0.98 | 0.96 | 0.97 | 4 | 0 | 238 | 0 |
|  |  |  | NORMAL | 0.9897 | 0.96 | 1.0 | 0.98 | 0 | 0 | 10 | 232 |

**Fig. 7.**
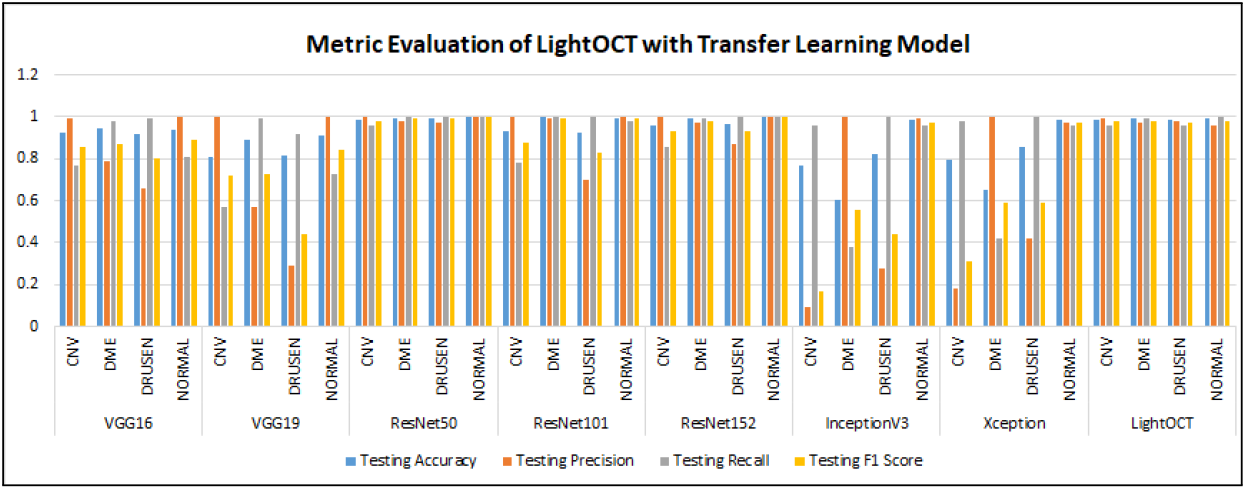
Metric evaluation of LightOCT with transfer learning models - VGG16, VGG19, ResNet50, ResNet101, InceptionV3 and Xception.

**Fig. 8.**
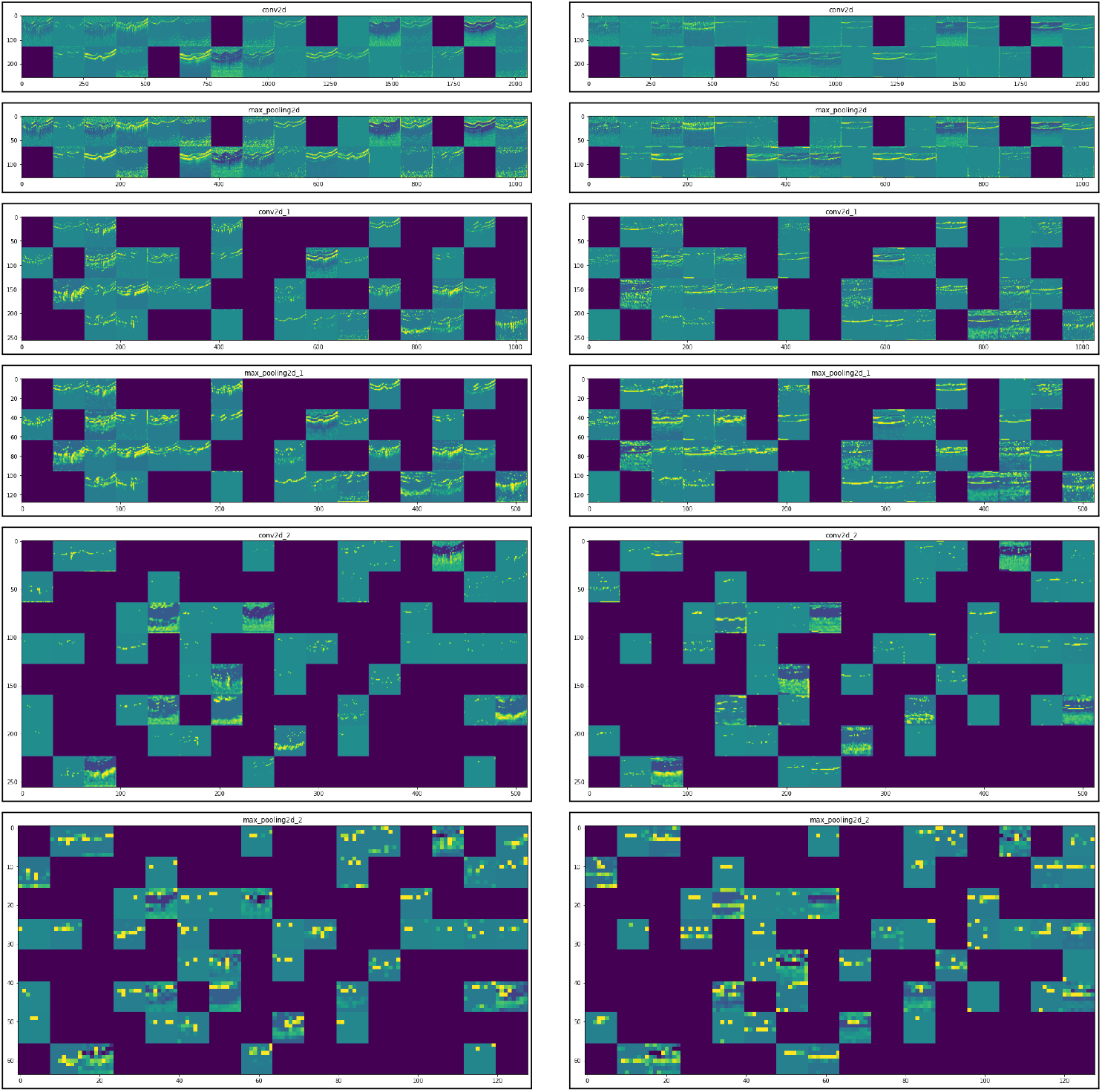
Results of neural Activation of CNN on (a) CNV and (b) DME

**Fig. 9.**
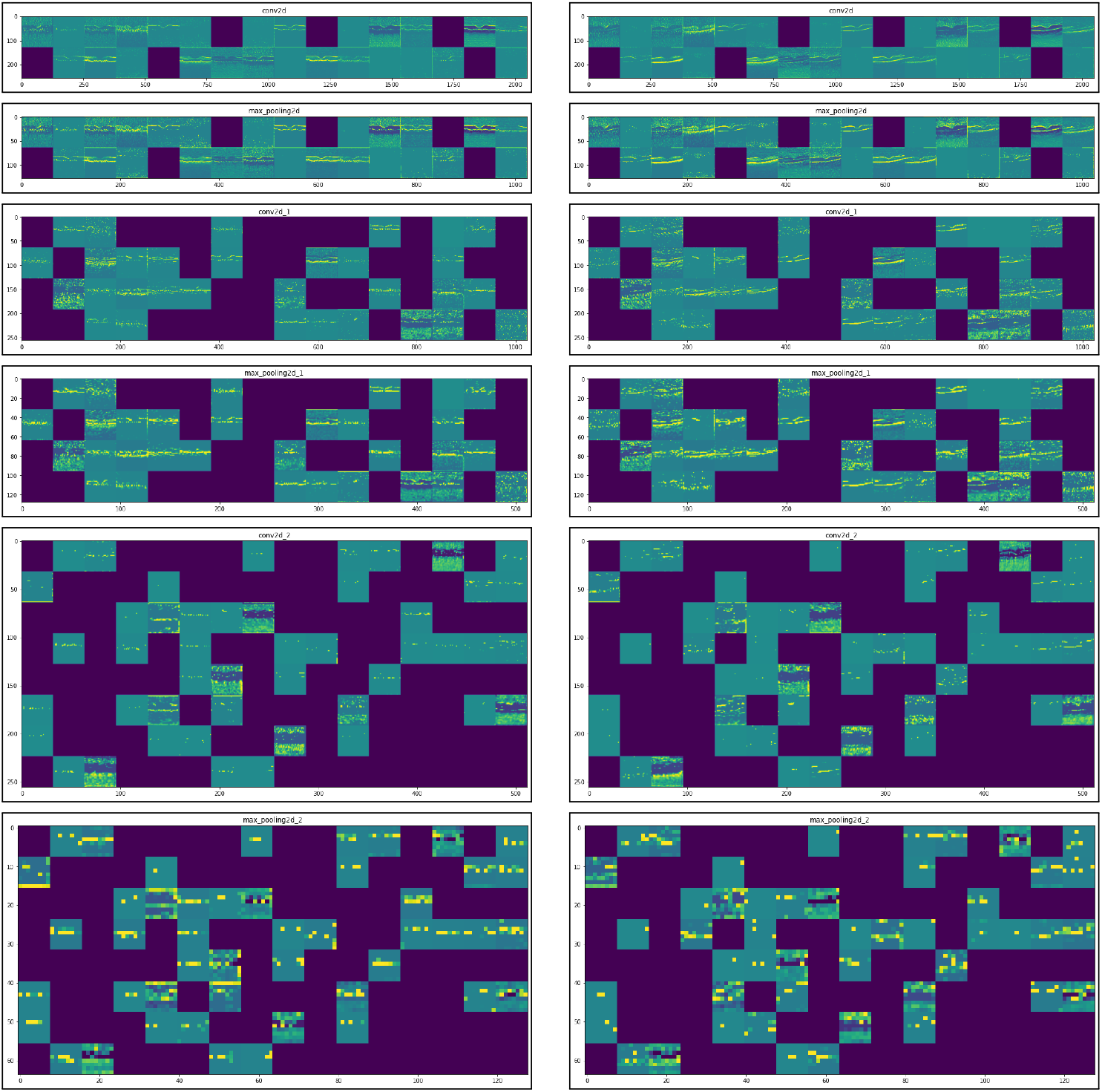
Results of neural Activation of CNN on (a) DRUSEN and (b) Normal

## 6 Discussion and Conclusion

In the present research, we examined the depth component in CNN architecture for retinal disease detection in Optical Coherence Tomography(OCT) images. Our results prove that shallow-feed forward neural networks can learn complex features and achieve comparable accuracies when compared with deep models, even on a small clinical dataset. In our analysis, we performed three experiments to validate our hypothesis. Firstly, we compare the architectures based on their depth, parameters, and regularization techniques used. The proposed architecture - LightOCT outperformed the other deep model configurations, with the lowest value of LVCEL as 0.2018 and the highest accuracy for each class. Secondly, we experimented to find the best fit optimizer for LightOCT. Even though Adam and RMSProp gave comparable results, the former exceeded RMSProp by an accuracy value differing a few decimal points. Finally, we chose the combination of LightOCT and Adam as the optimal architecture for our comparison. The last experiment was conducted on the comparative analysis of the state-of-the-art transfer learning models and our architecture. Even though ResNet variants outperformed LightOCT in the accuracy metric by some trailing decimal points, our proposed model is computationally efficient, and required the least training time, and gave comparable results with them. We would direct our future work to improve the accuracy metrics with shallow models such that the trade-off between training time and accuracy is reduced.

## Data Availability

All data produced are available online at: https://www.kaggle.com/paultimothymooney/kermany2018

## Notes

### Competing Interest Statement

The authors have declared no competing interest.

### Funding Statement

This study did not receive any funding

### Author Declarations

The present research used 84,495 optical coherence tomography (OCT) images collected by Kermany et al. in from various tertiary hospitals [Beijing Tongren Eye Center, California Retinal Research Foundation, Shiley Eye Institute of the University of California San Diego, Shanghai First People's Hospital, Medical Center Ophthalmology Associates] between July 1, 2013, and March 1, 2017. The data set has been clinically approved. Each OCT image was graded using a multi-tiered system. The initial grading round was completed by undergraduate and medical students who had completed an OCT review course. This group discarded any OCT images with artifacts or poor image resolution. The images passed on by the first group were approved or disapproved by the next tier of four ophthalmologists. They detected the presence or absence of various pathologies visible on the OCT based on their level of expertise. The truth labels for each image were validated by the final tier, which consisted of two retinal specialists with over 20 years of clinical experience. In addition, to account for human error in grading, a validation subset of 993 OCT images was verified by two additional ophthalmologist graders. The final dataset contains 37,205 class CNV, 11,348 showing DME, 8616 showing drusen, and 26,315 standard images. The test data has 242 images from each class. Link to data: https://www.kaggle.com/paultimothymooney/kermany2018

## References

[1] Lemaître, G., Rastgoo, M., Massich, J., Cheung, C.Y., Wong, T.Y., Lamoureux, E., Milea, D., Mériaudeau, F., Sidibé, D.: Classification of sd-oct volumes using local binary patterns: experimental validation for dme detection. Journal of ophthalmology 2016 (2016)

[2] Gulshan, V., Peng, L., Coram, M., Stumpe, M.C., Wu, D., Narayanaswamy, A., Venugopalan, S., Widner, K., Madams, T., Cuadros, J., et al.: Development and validation of a deep learning algorithm for detection of diabetic retinopathy in retinal fundus photographs. Jama 316(22), 2402–2410 (2016)

[3] Alsaih, K., Lemaitre, G., Rastgoo, M., Massich, J., Sidibé, D., Meriaudeau, F.: Machine learning techniques for diabetic macular edema (dme) classification on sd-oct images. Biomedical engineering online 16(1), 1–12 (2017)

[4] Karri, S.P.K., Chakraborty, D., Chatterjee, J.: Transfer learning based classification of optical coherence tomography images with diabetic macular edema and dry age-related macular degeneration. Biomedical optics express 8(2), 579–592 (2017)

[5] Hussain, M.A., Bhuiyan, A., D. Luu C.,, Theodore Smith, R., H. Guymer R.,, Ishikawa, H., S. Schuman J.,, Ramamohanarao, K.: Classification of healthy and diseased retina using sd-oct imaging and random forest algorithm. PloS one 13(6), 0198281 (2018)

[6] Srinivasan, P.P., Kim, L.A., Mettu, P.S., Cousins, S.W., Comer, G.M., Izatt, J.A., Farsiu, S.: Fully automated detection of diabetic macular edema and dry age-related macular degeneration from optical coherence tomography images. Biomedical optics express 5(10), 3568–3577 (2014)

[7] Tan, J.H., Bhandary, S.V., Sivaprasad, S., Hagiwara, Y., Bagchi, A., Raghavendra, U., Rao, A.K., Raju, B., Shetty, N.S., Gertych, A., et al.: Age-related macular degeneration detection using deep convolutional neural network. Future Generation Computer Systems 87, 127–135 (2018)

[8] Lu, W., Tong, Y., Yu, Y., Xing, Y., Chen, C., Shen, Y.: Deep learning-based automated classification of multi-categorical abnormalities from optical coherence tomography images. Translational vision science & technology 7(6), 41–41 (2018)

[9] Li, F., Chen, H., Liu, Z., Zhang, X., Wu, Z.: Fully automated detection of retinal disorders by image-based deep learning. Graefe’s Archive for Clinical and Experimental Ophthalmology 257(3), 495–505 (2019)

[10] Kermany, D.S., Goldbaum, M., Cai, W., Valentim, C.C., Liang, H., Baxter, S.L., McKeown, A., Yang, G., Wu, X., Yan, F., et al.: Identifying medical diagnoses and treatable diseases by image-based deep learning. Cell 172(5), 1122–1131 (2018)

[11] De Fauw, J., Ledsam, J.R., Romera-Paredes, B., Nikolov, S., Tomasev, N., Blackwell, S., Askham, H., Glorot, X., O’Donoghue, B., Visentin, D., et al.: Clinically applicable deep learning for diagnosis and referral in retinal disease. Nature medicine 24(9), 1342–1350 (2018)

[12] Ciregan, D., Meier, U., Schmidhuber, J.: Multi-column deep neural networks for image classification. In: 2012 IEEE Conference on Computer 18 LightOCT: Exploring the depth for Retinal disease detection Vision and Pattern Recognition, pp. 3642–3649 (2012). IEEE

[13] LeCun, Y., Boser, B., Denker, J.S., Henderson, D., Howard, R.E., Hubbard, W., Jackel, L.D.: Backpropagation applied to handwritten zip code recognition. Neural computation 1(4), 541–551 (1989)

[14] Khan, A., Sohail, A., Zahoora, U., Qureshi, A.S.: A survey of the recent architectures of deep convolutional neural networks. CoRR abs/1901.06032 (2019) 1901.06032

[15] Bouvrie, J.: 1 introduction notes on convolutional neural networks (2006)

[16] Ranzato, M., Huang, F.J., Boureau, Y.-L., LeCun, Y.: Unsupervised learning of invariant feature hierarchies with applications to object recognition. In: 2007 IEEE Conference on Computer Vision and Pattern Recognition, pp. 1–8 (2007). IEEE

[17] Szegedy, C., Liu, W., Jia, Y., Sermanet, P., Reed, S., Anguelov, D., Erhan, D., Vanhoucke, V., Rabinovich, A.: Going deeper with convolutions. In: Proceedings of the IEEE Conference on Computer Vision and Pattern Recognition, pp. 1–9 (2015)

[18] Kingma, D.P., Ba, J.: Adam: A method for stochastic optimization. arXiv preprint 1412.6980 (2014)

[19] Zeiler, M.D.: ADADELTA: an adaptive learning rate method. CoRR abs/1212.5701 (2012) 1212.5701

[20] Deng, J., Dong, W., Socher, R., Li, L.-J., Li, K., Fei-Fei, L.: Imagenet: A large-scale hierarchical image database. In: 2009 IEEE Conference on Computer Vision and Pattern Recognition, pp. 248–255 (2009). https://doi.org/10.1109/CVPR.2009.5206848

[21] Krizhevsky, A., Hinton, G., et al.: Learning multiple layers of features from tiny images (2009)

[22] Everingham, M., Van Gool, L., Williams, C.K.I., Winn, J., Zisserman, A.: The pascal visual object classes (voc) challenge. International Journal of Computer Vision 88(2), 303–338 (2010)

[23] Simonyan, K., Zisserman, A.: Very deep convolutional networks for largescale image recognition. arXiv preprint 1409.1556 (2014)

[24] He, K., Zhang, X., Ren, S., Sun, J.: Deep residual learning for image recognition. CoRR abs/1512.03385 (2015) 1512.03385

[25] Szegedy, C., Liu, W., Jia, Y., Sermanet, P., Reed, S.E., Anguelov, D., Erhan, D., Vanhoucke, V., Rabinovich, A.: Going deeper with convolutions. CoRR abs/1409.4842 (2014) 1409.4842

[26] Chollet, F.: Xception: Deep learning with depthwise separable convolutions. CoRR abs/1610.02357 (2016) 1610.02357

[27] Khan, A., Sohail, A., Zahoora, U., Qureshi, A.S.: A survey of the recent architectures of deep convolutional neural networks. Artificial Intelligence Review 53(8), 5455–5516 (2020)

[28] Kermany, D., Zhang, K., Goldbaum, M., et al.: Labeled optical coherence tomography (oct) and chest x-ray images for classification. Mendeley data 2(2) (2018)

